# Comparisons of Respiratory Syncytial Virus (RSV) and Influenza: Population Characteristics and Clinical Outcomes in Hospitalized Adults

**DOI:** 10.1101/2022.11.04.22281243

**Authors:** Katherine M. Begley, Arnold S. Monto, Lois E. Lamerato, Anurag N. Malani, Adam S. Lauring, H. Keipp Talbot, Manjusha Gaglani, Tresa McNeal, Fernanda P. Silveira, Richard K. Zimmerman, Donald B. Middleton, Shekhar Ghamande, Kempapura Murthy, Lindsay Kim, Jill M. Ferdinands, Manish M. Patel, Emily T. Martin

**Affiliations:** Department of Epidemiology, University of Michigan School of Public Health, Ann Arbor, Michigan, USA; Department of Public Health Sciences, Henry Ford Health, Detroit, Michigan, USA; Department of Medicine, Section of Infectious Diseases, St Joseph Mercy Hospital, Ann Arbor, Michigan, USA; Department of Infection Prevention and Control, St Joseph Mercy Hospital, Ann Arbor, Michigan, USA; Department of Internal Medicine, University of Michigan Medical School, Ann Arbor, Michigan, USA; Department of Medicine and Health Policy, Vanderbilt University Medical Center, Nashville, Tennessee, USA; Departments of Pediatrics, Section of Pediatric Infectious Diseases (MG), Internal Medicine, Section of Hospital Medicine (TM), Section of Critical Care and Pulmonary Medicine (SK) and Data/Biostatistics Research Core (KM), Baylor Scott & White Health, Temple, Texas, USA; Department of Medical Education at Texas A&M University COM, Temple, Texas, USA; University of Pittsburgh and University of Pittsburgh Medical Center, Pittsburgh, Pennsylvania, USA; Division of Viral Diseases, Centers for Disease Control and Prevention, Atlanta, Georgia, USA; US Public Health Service, Rockville, Maryland, USA; Influenza Division, Centers for Disease Control and Prevention, Atlanta, Georgia, USA

## Abstract

**Background:** Respiratory syncytial virus (RSV) is under-recognized in hospitalized adults. We evaluated severity of acute respiratory illness (ARI) including intensive care unit (ICU) admission and mechanical ventilation in a national surveillance network.

**Methods:** Hospitalized adults who met a standardized ARI case definition were prospectively enrolled across three respiratory seasons from hospitals participating across all sites of the U.S. Hospitalized Adult Influenza Vaccine Effectiveness Network (HAIVEN, 2016-2019). Multivariable logistic regression was used to test associations between lab-confirmed infection and characteristics and clinical outcomes.

**Results:** Among 10,311 hospitalized adults, 6% tested positive for RSV (n=622), 18.8% positive for influenza (n=1,940), and 75.1% negative for RSV and influenza (n=7,749). The proportion of adults with Congestive Heart Failure (CHF) or Chronic Obstructive Pulmonary Disease (COPD) was higher among adults with RSV than influenza (CHF: 37.3% vs. 28.8%, p<0.0001; COPD: 47.6% vs. 35.8%, p<0.0001). Patients with RSV had higher odds of experiencing length of stay ≥8 days [OR=1.38 (95% CI: 1.06-1.80), p-value=0.02] and invasive or noninvasive mechanical ventilation [OR=1.45 (95% CI: 1.09-1.93), p-value=0.01] compared with influenza patients.

**Conclusions:** Our findings suggest patients with RSV might incur worse outcomes than influenza in hospitalized adults, who are likely to have pre-existing cardiopulmonary conditions.

## Introduction

Respiratory syncytial virus (RSV) is widely regarded as a disease of young children most severe in infants < 2 years [1,2]. However, clinically significant RSV infection occurs at all ages, and adults may present with illness ranging from mild upper respiratory infections to severe respiratory distress [3,4]. Prior research has identified the following patient risk factors for RSV associated hospitalization: severe immunocompromising conditions, underlying cardiopulmonary comorbidities including congestive heart failure (CHF) and chronic obstructive pulmonary disease (COPD), and age with frailty [5–10]. Additional research is needed to support the ongoing evaluation of RSV hospitalizations in adults. Moreover, results from such research have the potential to inform targeted vaccination strategies to protect high-risk adults from adverse health outcomes once a safe and effective vaccine is available.

Hospitalizations and in-hospitalization outcomes, such as intensive care unit (ICU) admission and the need for mechanical ventilation, are widely accepted as markers of clinical severity with respect to acute respiratory illness (ARI). Moreover, hospitalization with influenza is well-documented and well-researched, making it a strong benchmark for comparing RSV hospitalization outcomes [11,12,12,14]). A prior analysis of one Hospitalized Adult Influenza Vaccine Effectiveness Network (HAIVEN) site (Michigan) concluded patients hospitalized with RSV, compared to patients hospitalized with influenza, had more comorbidities and a longer symptomatic period prior to hospital admission (days) [13]).

This study aims to characterize the frequency and clinical severity of RSV among hospitalized adults ≥ 18 years for three respiratory illness seasons (2016-2019) across surveillance sites in four states participating in the HAIVEN study. This study has two objectives: first, to identify population characteristics and key differences among adults (ages ≥18 years) and, second, to compare clinical outcomes among adults hospitalized with RSV, influenza, or neither from all sites participating in the HAIVEN study.

## Methods

### Source Population

Data collected for this analysis comes from the HAIVEN study — a prospective study of adults hospitalized with ARI meeting a standardized case definition [12,13,15]. The HAIVEN study was designed as a lab-confirmed case - test negative control study aimed at estimating vaccine effectiveness in the prevention of hospitalization associated with adult influenza cases [12,13,15]. Adults ≥18 years old admitted to a participating HAIVEN site hospital were prospectively identified from September 2016 through May 2019 through either chief complaint(s) and/or admission diagnosis of an ARI. HAIVEN is comprised of eight hospitals in 2015-2016, nine in 2016-2017, and ten in 2017-2019, with locations in Michigan, Pennsylvania, Texas, and Tennessee [16]. Participating sites included both academic medical centers and community hospitals. HAIVEN study eligibility criteria have been described elsewhere [12,13,15,16]. To participate in the study, written informed consent was provided by patients or a proxy/surrogate. This protocol was reviewed and approved by the University of Michigan Institutional Review Board, the Vanderbilt University Medical Center Institutional Review Board, the University of Pittsburgh Human Research Protection Office, and the Baylor Scott & White Research Institute Institutional Review Board. The Centers for Disease Control and Prevention Institutional Review Board issued a reliance on the site specific approvals.

### Data Collection

Through structured enrollment interviews with HAIVEN research staff, consenting participants self-reported, or reported via proxy/surrogate where appropriate, demographic data, illness onset date, frailty score (0, 1, 2, 3, 4/5), and current and prior season influenza vaccination status (yes/no). At the time of the enrollment interview, research staff collected throat and nasal swabs, which were combined in universal transport media (UTM) or clinical mid-turbinate swabs were tested with either singleplex or multiplex respiratory pathogen panel molecular assays per site protocol. Specimens were transported to HAIVEN site laboratories and tested for RSV and influenza using real-time reverse transcription PCR (RT-PCR) with primers, probes, and protocols developed by the CDC Division of Viral Diseases and Influenza Division or with existing clinical laboratory assays following completion of CDC proficiency testing. Electronic medical records (EMR) were reviewed to extract data for calculating Charlson Comorbidity Index (CCI) scores (0, 1-2, and ≥ 3), determining body mass index (BMI), as well as documented evidence of COPD, CHF, and asthma. Obesity was defined as having a BMI ≥ 30. The outcomes of interest were also extracted from participant EMR including length of stay, ICU admission, need for invasive or non-invasive mechanical ventilation, and death prior to or 30 days after discharge. An extended length of stay was defined as ≥8 days.

### Statistical Analysis

Descriptive statistics were calculated for the following variables across each comparison group of interest: age group (18-49, 50-64, and 65+ years), sex, race/ethnicity (White non-Hispanic, Black non-Hispanic, Other non-Hispanic, and Hispanic), BMI (normal, overweight, and obese), CCI scores, asthma, CHF, COPD, frailty, site (Michigan, Texas, Pennsylvania, and Tennessee), season (2016/17, 2017/18, and 2018/19), and influenza vaccination status. Descriptive statistics were calculated for in-hospital outcomes of interest across each comparison group of interest, including extended length of stay, ICU admission, mechanical ventilation (any), and death (pre-discharge and 30 days post-discharge). CCI scores, time from illness onset to hospital admission, and time from illness onset to lab specimen collection were described with median and interquartile range (IQR) statistics. CCI scores of individuals who tested positive for RSV were compared to individuals who tested positive for influenza using Wilcoxon rank-sum tests, stratified by age group (18-49, 50-64, and 65+ years). Overall and age-stratified (18-49, 50-64, and 65+ years) proportions of CHF and COPD for RSV-positive participants were compared to proportions of CHF and COPD for influenza-positive participants using Chi-square statistics. Age-adjusted, Firth logistic regression models were used to test the association between CHF/COPD and RSV detection compared to influenza detection.

We evaluated characteristics and clinical outcomes by comparing RSV-positive versus two comparison groups separately: 1) influenza-positive; and 2) RSV-negative/influenza-negative cases. Multivariable logistic regression models were used to assess participant characteristics as risk factors – age, sex, CCI, BMI, site, season, time from illness onset to admission, and time from illness onset to specimen collection – associated with case detection status using the two comparison groups. Multivariable logistic regression models were used to test the association between clinical outcomes of interest – extended length of stay, ICU admission, need for any mechanical ventilation, and death – and case detection status using the above comparison groups. Participant characteristics and clinical outcome multivariable models were adjusted for age, race/ethnicity, sex, BMI, CCI, site, season, and time from illness onset to admission. To account for small cell counts resulting from stratification, all logistic models used Firth’s adjustment [17]. Profile-likelihood confidence intervals and Wald p-values were used to determine statistical significance. A p-value less than 0.05 was considered statistically significant for all analyses, and all analyses were performed using SAS software version 9.4 (SAS Institute, Cary, NC).

## Results

### Population Characteristics & RSV-Influenza Epidemiology

11,369 adults were enrolled in HAIVEN across the three respiratory seasons included in this study. Individuals with subsequent enrollments within a given respiratory illness season (n=602) and/or with missing influenza or RSV laboratory results (n=499), and those presenting with an RSV-influenza co-infection (n=21) were excluded from analysis. The final study population for this analysis was comprised of 10,311 inpatients, 26.0% (n=2,679) from 2016-2017, 37.7% (n=3,885) from 2017-2018, and 36.3% (n=3,747) from 2018-2019 (Table 1).

**Table 1.**
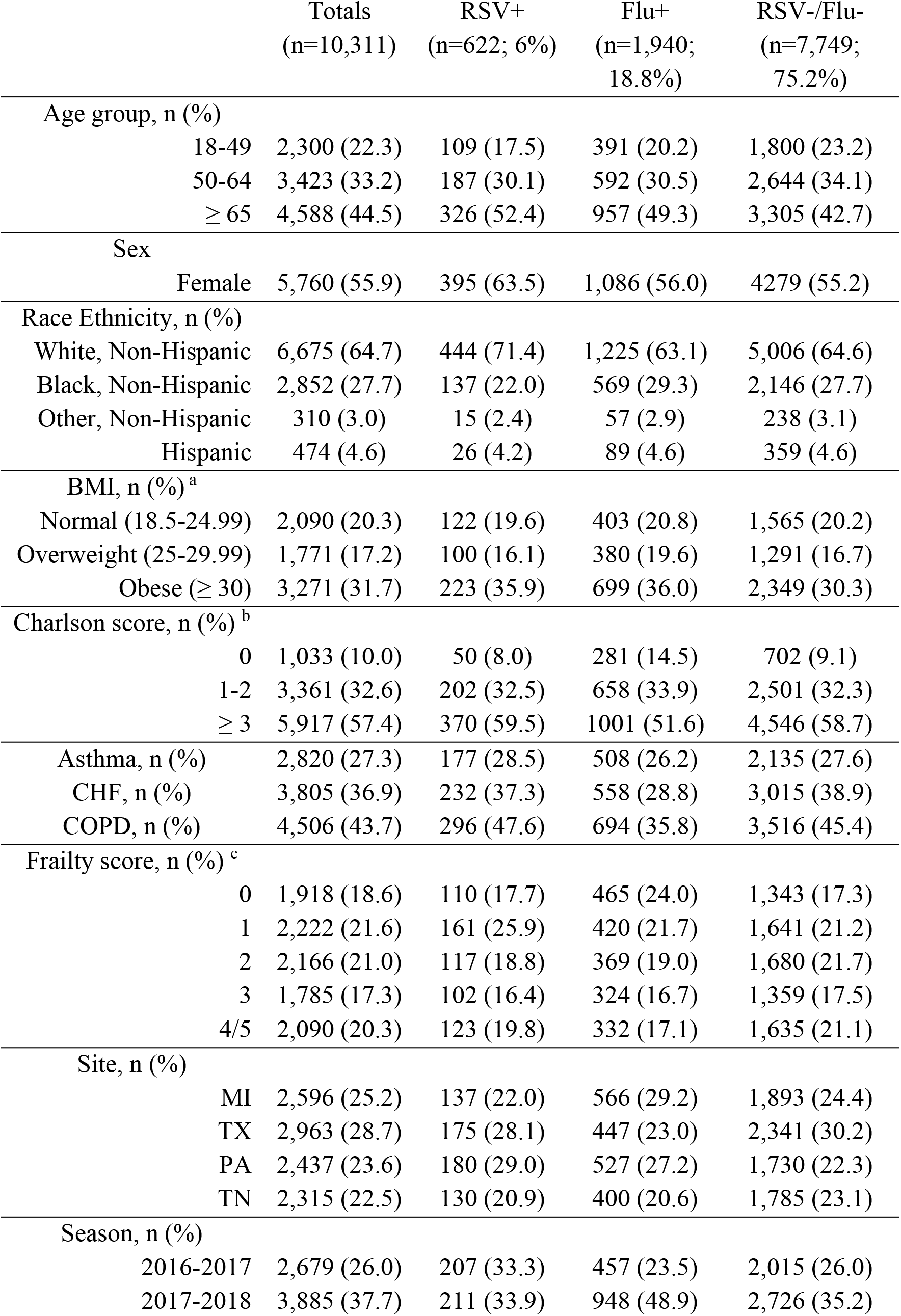
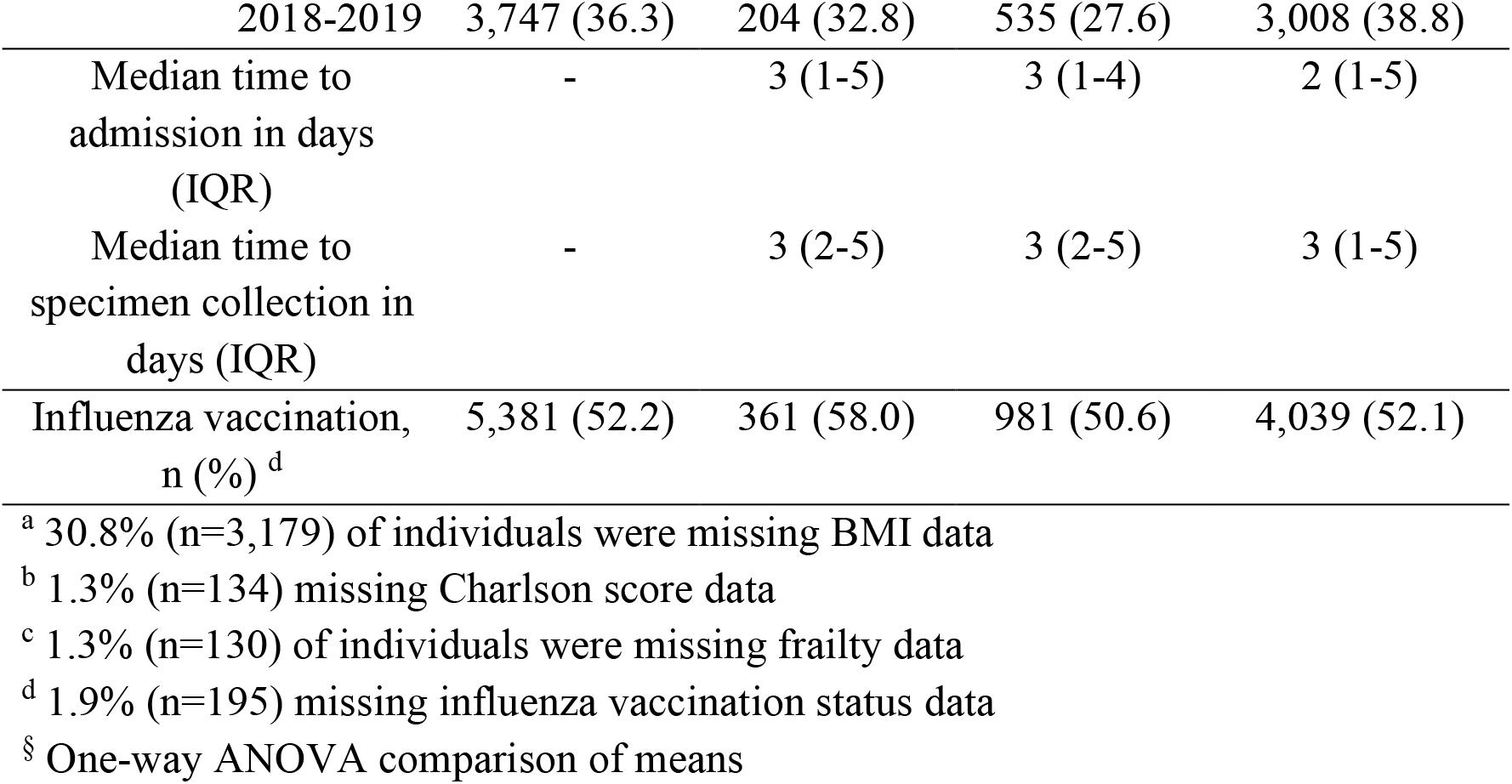
Epidemiologic Characteristics Among Participants

Overall, RSV was detected in 6.0% (n=622) and influenza in 18.8% (n=1,940) of eligible participants. The remaining 75.2% (n=7,749) of included participants had neither RSV nor influenza detected. RSV peaked in January for the 2016-2017 and 2017-2018 seasons but in December in 2018-2019 (Figure 1). On the other hand, the peak for influenza tended to vary with peaks occurring in February, January, and March, respective to each season included in this analysis.

**Figure 1.**
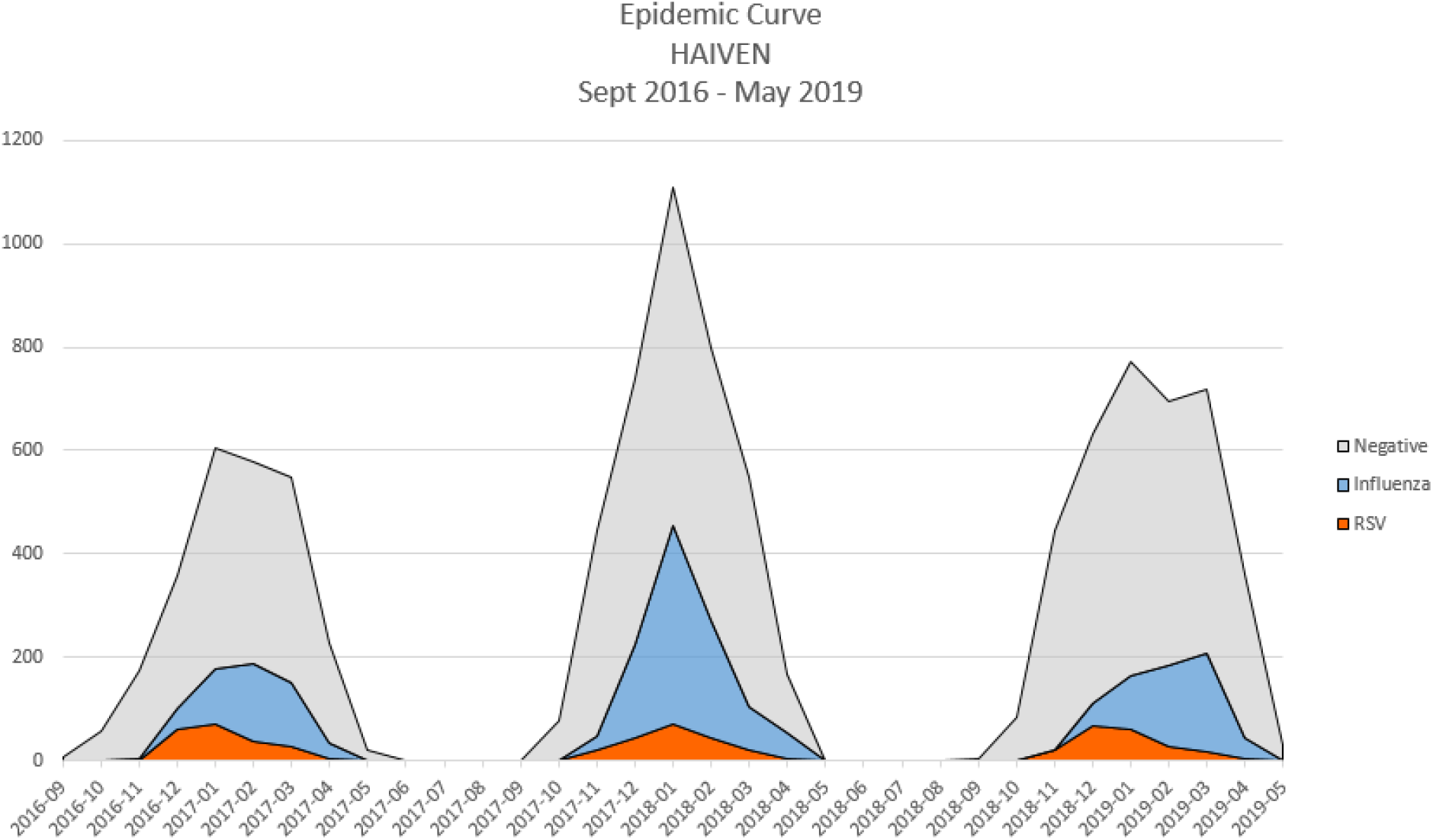
Modified HAIVEN RSV-Influenza Epidemic Curve.

Just over half (52.4%) of all RSV cases in this study were among adults ≥ 65 years of age (Table 1). The age-specific proportion of RSV among adults hospitalized with ARI increased within each age group was 4.7% among those 18-49, 5.5% among those 50-64, and 7.1% among those ≥65 years of age. The age-specific prevalence of influenza was relatively consistent among those aged 18-49 (17%) and 50-64 (17.3%) with slightly higher (20.9%) detection among those ≥65 years of age. The median time from illness onset to hospital admission among RSV-positive patients was 3 days (IQR: 1-5), and similar to that among influenza-positive patients was 3 days (IQR: 1-4). Median time from illness onset to specimen collection for patients with influenza or RSV infection was 3 days (IQR: 2-5) (Table 1).

Overall, 90% (n=9,278) of included participants had a CCI ≥1, indicating the presence of at least one underlying chronic comorbid condition (Table 1), and this finding was expected given all patients were hospitalized. The overall proportion of adults with CHF or COPD was significantly (CHF p<0.0001; COPD p<0.0001; *X*^*2*^ test) higher in those with RSV (37.3% CHF, 47.6% COPD) compared with those with influenza (28.8% CHF, 35.8% COPD) (Table 1). When stratified by age group, among adults aged 50-64 years, the proportions of CHF and COPD between RSV-positive (31.0% CHF, 47.1% COPD) and influenza-positive (25.0% CHF, 43.4% COPD) patients were comparable (CHF p=0.10; COPD p=0.38; *X*^*2*^ test). Among adults aged 18-49 years, RSV-positive patients had higher proportions of CHF and COPD (26.6% CHF, 29.4% COPD) when compared to influenza-positive patients (14.3% CHF, 13.6% COPD) (CHF p=0.003; COPD p=0.0001; *X*^*2*^ test). In adults aged 65 years and older, those with RSV had higher proportions of CHF and COPD (44.5% CHF, 54.0% COPD) when compared to those with influenza (37.0% CHF, 40.1% COPD) (CHF p=0.02; COPD p<0.0001; *X*^*2*^ test). Adjusting for age, when compared to inpatients without CHF, those with CHF had significantly higher odds of having RSV compared to influenza [OR_adj_=1.40 (95% CI: 1.15-1.70), p-value=0.0007]. Adjusting for age, when compared to inpatients without COPD, those with COPD had significantly higher odds of having RSV compared to influenza [OR_adj_=1.57 (95% CI: 1.31-1.89), p-value<0.0001]. In the age groups of 18-49 and ≥65 years, those with RSV had significantly higher median CCI scores of 2 (1-4) and 4 (2-6) when compared to those with influenza of 1(1-3) and 3(1-6), respectively (p=0.02 each) (Table 2, Figure 2).

**Table 2.**
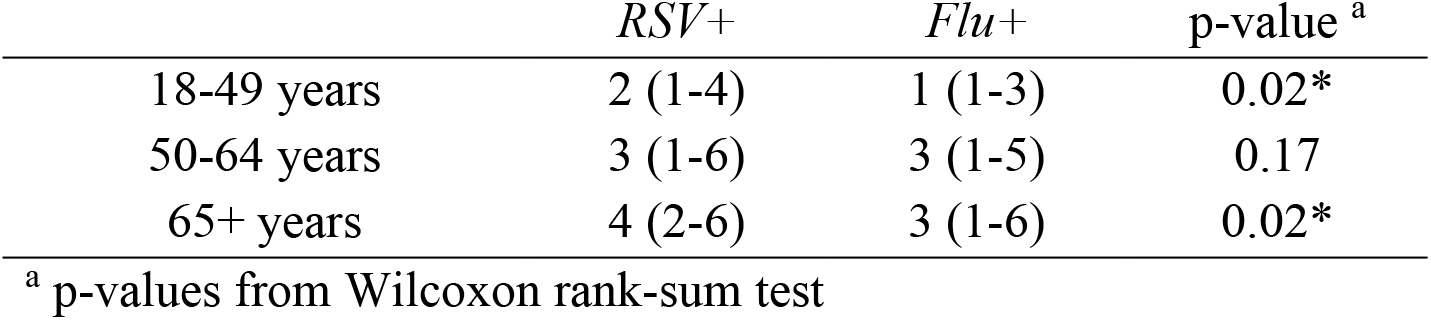
Comparison of median (IQR) CCI among RSV-positive and influenza-positive hospitalizations, stratified by age-group

**Figure 2.**
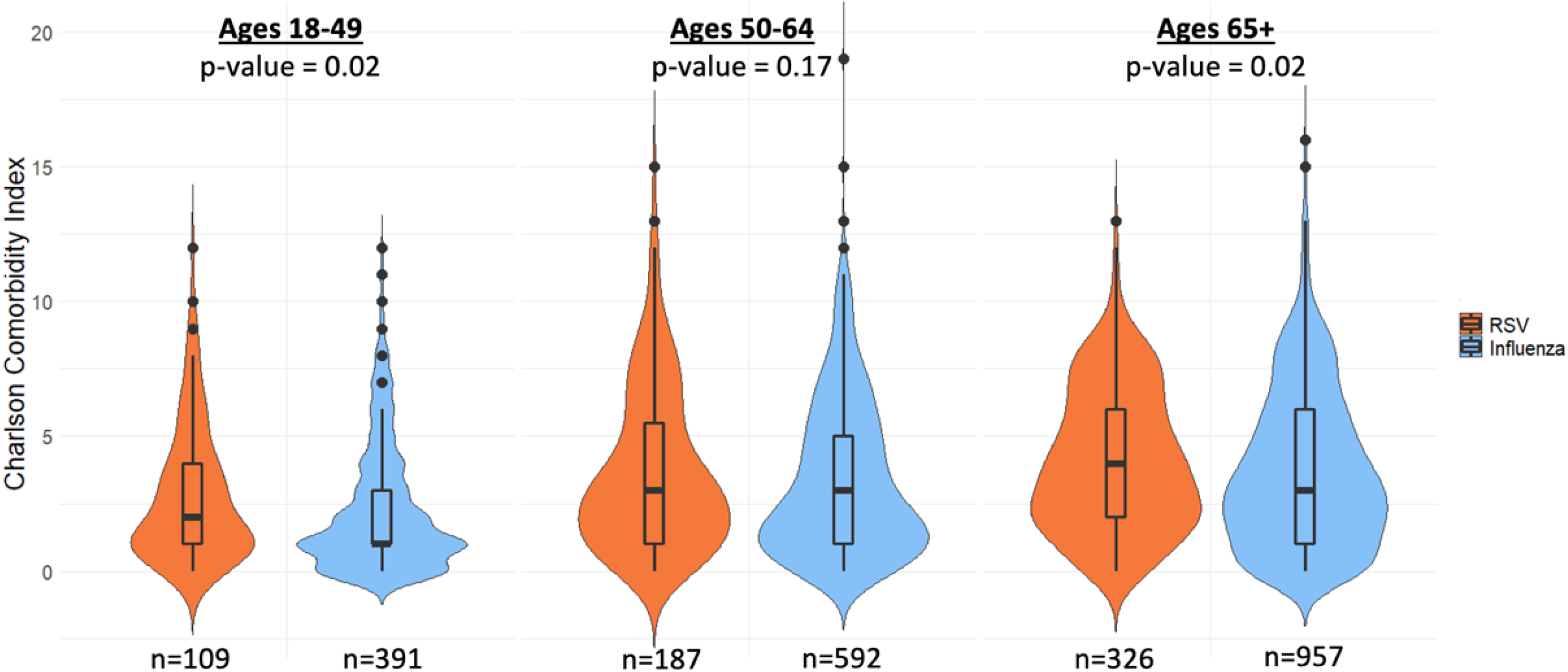
Violin Plot Comparing RSV and Influenza Participant CCI Stratified by Age.

### Clinical Outcomes

Among RSV-positive inpatients, 16.6% experienced a length of hospital stay ≥8 days, 12.4% were admitted to the ICU, and 15% required some type of mechanical ventilation — either invasive or noninvasive (Table 3). In contrast, among patients with influenza, 11.3% experienced a length of stay ≥8 days, 9.9% were admitted to the ICU, and 11.1% required some type of mechanical ventilation (Table 3). With respect to patient deaths captured by EHR, nine (1.5%) occurred in RSV-positive patients, 25 (1.3%) in patients with influenza, and 105 (1.4%) in the RSV and influenza test negative group (Table 3).

**Table 3.** Frequencies and Odds Ratios of Clinical Outcomes Comparing Hospitalized ARI Adults, (n) observations overall and included in model

| *Frequencies of Outcomes* | *n (%)* |  |  |  |
| --- | --- | --- | --- | --- |
|  | Total<br>(n=10,311) | RSV<br>(n=622) | Influenza<br>(n=1,940) | RSV-/Flu-<br>(n=7,749) |
| Extended LOS ($\geq 8$ days) | 1,616 (15.7) | 103 (16.6) | 220 (11.3) | 1,293 (16.7) |
| ICU Admission <sup>x</sup> | 1,340 (13.0) | 77 (12.4) | 192 (9.9) | 1,071 (13.8) |
| Mechanical ventilation, any <sup>x</sup> | 1,447 (14.0) | 93 (15.0) | 215 (11.1) | 1,139 (14.7) |
| Death |  |  |  |  |
| During hospitalization <sup>§</sup> | 139 (1.4) | 9 (1.5) | 25 (1.3) | 105 (1.4) |
| 30 days post-discharge <sup>†</sup> | 222 (2.2) | 11 (1.8) | 23 (1.2) | 188 (2.4) |
| *Adjusted logistic regression models OR (95% CI) <sup>a</sup>* |  |  |  |  |
|  | RSV vs.<br>Influenza<br>(n=2562) | p-value <sup>b</sup> | RSV vs.<br>RSV-/Flu-<br>(n=8371) | p-value <sup>b</sup> |
| Extended LOS ($\geq 8$ days) | 1.40<br>(1.08-1.82) | 0.01* | 1.03<br>(0.82-1.28) | 0.83 |
| ICU Admission <sup>x</sup> | (n=2558) |  | (n=8359) |  |
|  | 1.27<br>(0.95-1.69) | 0.11 | 0.95<br>(0.74-1.22) | 0.70 |
| Mechanical ventilation, any <sup>x</sup> | (n=2558) |  | (n=8359) |  |
|  | 1.46<br>(1.09-1.94) | 0.01* | 1.17<br>(0.91-1.49) | 0.21 |
| Death |  |  |  |  |
| During hospitalization | (n=2537) |  | (n=8325) |  |
|  | 0.94<br>(0.46-1.94) | 0.88 | 1.19<br>(0.61-2.31) | 0.61 |
| 30 days post-discharge | (n=1863) |  | (n=6516) |  |
|  | 1.36<br>(0.65-2.84) | 0.42 | 0.72<br>(0.38-1.37) | 0.32 |

<sup>x</sup> 0.14% (n=14) individuals missing ICU and mechanical ventilation data
<sup>§</sup> 0.6% (n=63) individuals missing inpatient death data
<sup>†</sup> 23.0% (n=2376) missing death after discharge data
<sup>a</sup> Adjusted for age, sex, race, Charlson score, BMI, site, season, and days to admission
<sup>b</sup> Wald p-values

After adjusting for potential confounders, when compared to influenza-positive inpatients, RSV-positive patients had significantly higher odds of experiencing an extended length of stay ≥8 days [OR_adj_=1.40 (95% CI: 1.08-1.82), p-value=0.01] and need for mechanical ventilation [OR_adj_=1.46 (95% CI: 1.09-1.94), p-value=0.01] (Table 3). There were no significant death or other clinical outcome associations when comparing inpatients with RSV to the RSV-negative, influenza-negative group.

### Participant Characteristics as Risk Factors

Patients with lab-confirmed RSV had twice the odds of having a CCI ≥ 3 compared with patients with influenza, which was statistically significant [OR_adj_=2.10 (95% CI: 1.50-2.93), p-value<0.0001] (Table 4). Obesity [OR_adj_=1.29 (95% CI: 1.02-1.63), p-value=0.03] and age ≥65 years [OR_adj_=1.65 (95% CI: 1.30-2.10), p-value<0.0001] were significantly associated with RSV detection when compared to participants negative for both RSV and influenza. Female sex was significantly associated with RSV detection when compared to patients with influenza [OR_adj_=1.44 (95% CI: 1.19-1.75), p-value=0.0002] as well as RSV-negative, influenza-negative [OR_adj_=1.38 (95% CI: 1.16-1.63), p-value=0.0003].

**Table 4.**
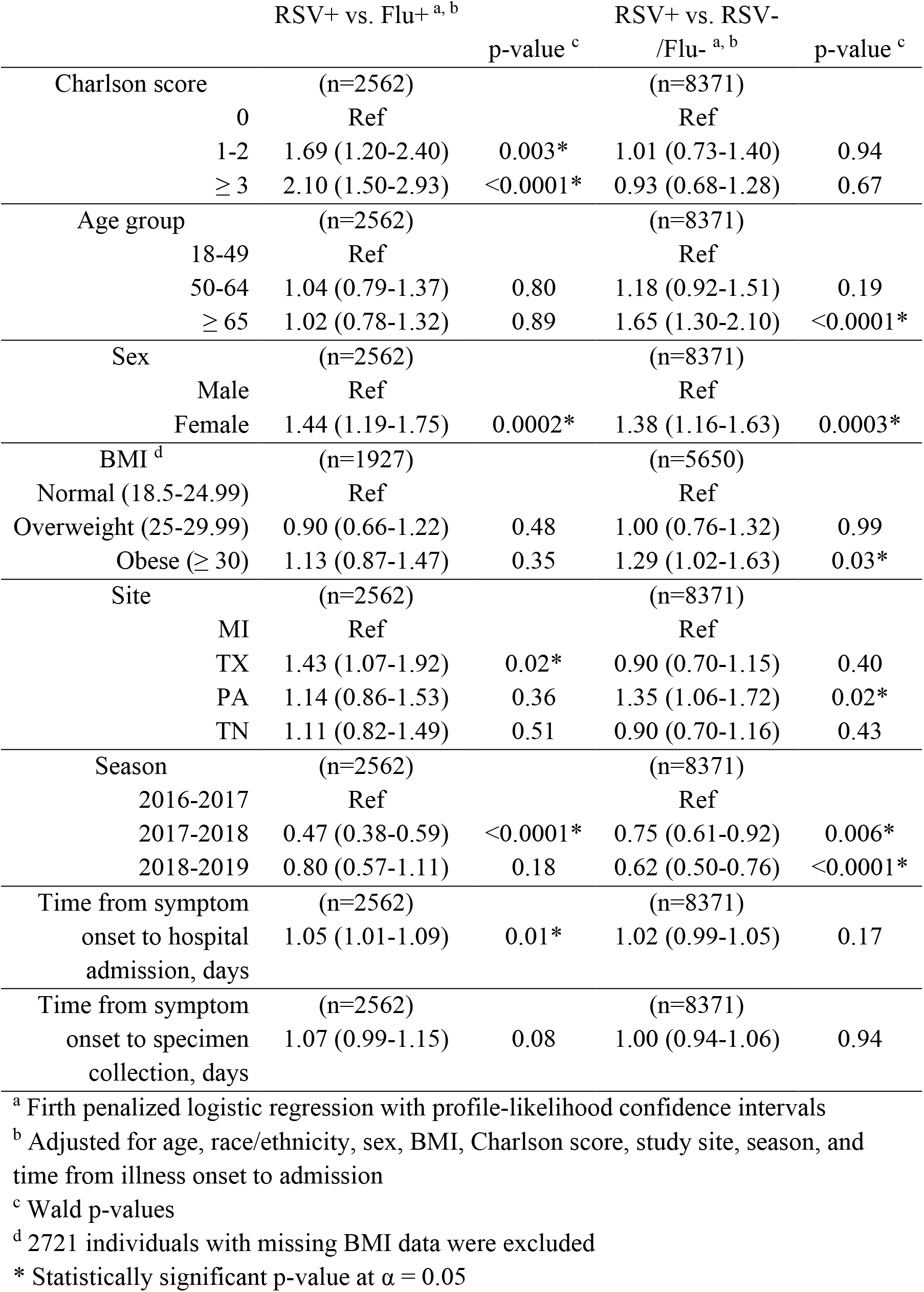
Factors associated with case detection using adjusted logistic regression models, OR (95% CI), (n) observations included in model

## Discussion

The goal of this study was to compare characteristics and clinical outcomes of adults hospitalized with lab-confirmed RSV versus influenza and RSV versus neither RSV nor influenza between 2016 and 2019 from hospitals at four sites participating in the HAIVEN study. Hospitalized adults with RSV had a greater overall proportion of underlying cardiopulmonary comorbidities and higher CCI scores when compared to those with influenza. With respect to clinical outcomes, hospitalized adults with RSV had higher odds of experiencing an extended length of hospital stay of 8 or more days and need for invasive or non-invasive mechanical ventilation when compared to those with influenza. Female sex and a slight increase in time from illness onset to hospital admission were associated with RSV when compared to those with influenza. Female sex, age 65 and older, and obesity were also associated with RSV when compared to those with neither influenza nor RSV.

Adults aged 18-49 and 65+ with RSV had significantly higher median CCI scores when compared to those with influenza, and the proportion of adults with CHF or COPD was significantly higher in those with RSV compared with those with influenza. Findings related to CCI in our study are consistent with the findings of Malosh et al [18]. Reproducibility in a larger, geographically diverse population is essential for strengthening criteria to identify adults who should be considered at high-risk of experiencing severe illness when infected with RSV. Furthermore, our findings corroborate studies that identified adults with a history of CHF and/or COPD as a high-risk group for infection with RSV [7,10].

Results from our clinical outcomes analysis indicate that RSV-associated ARI may be more severe than influenza-associated ARI in some instances. Fifty percent of adults with influenza detected had received an influenza vaccine, which may offer protection against severe influenza-associated outcomes. Furthermore, many adults hospitalized with influenza receive antiviral treatments that are not available for RSV. However, there were no differences in clinical outcomes when comparing the RSV-positive group to the RSV-negative/influenza-negative group, which may be due to these two groups being more similar, especially with regards to proportions of CHF and COPD detected. We found some differences in the demographics of the hospitalized RSV population as well. When compared to the RSV and influenza negative control group, those with RSV were more likely to be obese, female, and aged 65 and older. This result is consistent with the findings from Malosh et al.’s comparison of RSV and RSV-negative, influenza-negative patients [18]. Interestingly, the association measured among those aged ≥65 is unique, albeit expected, and may have been statistically significant due to the expanded sample size of our study.

When compared to each of those with influenza detected as well as RSV-negative, influenza-negative, female sex was significantly associated with RSV detection. A study of 2,225 adults aged ≥50 with medically attended ARI reported a null association between RSV detection and sex from adjusted logistic regression models [19]. Our finding could be in part due to a larger sample size as well as the inclusion of adults aged 18 years and older. While we do not have data on whether or not our participants work with children or have children residing in their household, it is possible that women are at higher risk of contracting RSV, considering they disproportionally assume roles with greater time spent among children, both personally and professionally. Alternatively, prior research suggests that biologic sex – through various mechanisms including environmental factors such as close contact with young children in the household as well as the presence of underlying chronic conditions – plays a key role in differential incidence, immune response, and differences in severity of ARI [20–23].

The analysis represents a large sample size with considerable geographic diversity due to the inclusion of all sites as well as more recent seasons of the HAIVEN study. Compared to a six-year retrospective study of adults hospitalized with RSV conducted in one U.S. city, we captured 133 more cases of RSV from the three seasons included in our analysis [24]. In the future, this study could be improved by including influenza and RSV viral subtype data to explore their impact on in-hospital outcomes. Another strength of using the HAIVEN study is the implementation of prospective, active participant enrollment that does not solely depend on clinical test orders for case status determination [12]. Missing data was not a restrictive analytic issue due to data collection from participant EMR; however, death post-discharge was missing for a considerable number of observations, which is expected given the potential for loss-to-follow-up in large, hospital-based studies, limiting our analysis of that outcome. Generalizability of this study is limited, and precaution should be taken when interpreting these results for populations that are not predominantly non-Hispanic.

Multiple RSV vaccine candidates in various stages of clinical trials have demonstrated promising results of safety and efficacy. Determining which populations to focus on during initial vaccine rollout is a high-priority discussion that should happen prior to availability of a vaccine. Yamin et al. posit that targeting children <5 years as a vaccine strategy would be the most efficient method of reducing RSV cases in children as well as adults and the elderly through indirect protection [25]. In a review by Stephens and Varga, the authors emphasize the need for a vaccine tailored to elderly populations that will elicit a high immune response to overcome immune dysfunction often associated with aging [26]. Our analysis supports previous research through the identification of adults with underlying cardiopulmonary comorbidities being at high risk of experiencing severe RSV illness. To reduce medical costs and resource burden associated with RSV hospitalization, consideration should be given to the immunocompromised, adults aged ≥65 years, and young children < 5 years in any targeted vaccination campaign.

## Data Availability

All data produced in the present study may be requested from study authors and the Centers for Disease Control and Prevention.

